# Probing the Biological Underpinnings of Advanced Brain Ageing in Schizophrenia

**DOI:** 10.1101/2024.07.05.24309965

**Authors:** Alex J. Murray, Pedro L. Ballester, Jack C. Rogers, Stephen Williams, Martin Wilson, Bill Deakin, Mohammad Katshu, Peter Liddle, Lena Palaniyappan, Rachel Upthegrove

## Abstract

**Background:** Recent evidence suggests that patients with schizophrenia may show advanced brain ageing, particularly evident after the first year of onset. However, it is unclear if accelerated ageing relents, persists or continues to increase over time. The underlying causal factors are also poorly understood. Disruptions in glutamate function, oxidative stress, and inflammation may all contribute to progressive brain changes in people with schizophrenia. We examine whether brain ageing differs between early and established stages of schizophrenia, correlates with symptom severity and varies with markers of brain function, oxidative status and inflammatory burden.

**Methods:** Two brain-age prediction models assessed 112 participants (34 recent onset psychosis, 36 established schizophrenia, 42 healthy controls). Brain age gap (BAG) was calculated by subtracting chronological age from predicted age. Shapley’s additive explanations (SHAP) identified influential structural magnetic resonance imaging (MRI) features driving brain-age prediction. Linear regression models and partial correlations, adjusting for age, explored associations between BAG and neurometabolites, inflammatory markers, medication exposure and clinical scores in the whole sample.

**Results:** The established schizophrenia group showed higher BAG (Mean = 6.21, SD = 7.30) compared to healthy individuals (Mean = -0.01, SD = 9.10), while recent-onset patients (Mean = 4.23, SD = 9.25) did not differ significantly from healthy individuals. The top 10 SHAP features diving the BAG included ventricular enlargement and total grey matter volume, which was similar in psychosis to healthy individuals. In a combined psychosis group (established + recent-onset), higher BAG correlated with more severe symptoms (PANSS total, general, and anxiodepressive subscales). BAG positively associated with Magnetic Resonance Spectroscopy measured glutathione and negatively with N-Acetyl Aspartate.

**Discussion:** Accelerated brain age in schizophrenia may be related to illness severity and poor defence against oxidative stress. The lack of differences in SHAP features between schizophrenia and healthy individuals suggests that the pattern of brain ageing is in keeping with advanced normal ageing. Findings suggest potential treatment targets to improve brain health in schizophrenia, warranting further research.

## 1 Introduction

Advanced brain-ageing (ABA) has been observed across various psychiatric disorders, with some evidence that these are present even in early stages (Koutsouleris et al., 2014; Schnack et al., 2016; Ballester et al., 2021). The quantification of ABA involves assessing the deviation between an individual’s actual age and a prediction derived from models trained on a large datasets of structural neuroimaging features from healthy individuals (Franke and Gaser, 2019; Sanford et al., 2022). Specifically in schizophrenia, the predicted acceleration in ageing is thought to be primarily driven by reduced grey matter tissue (Ballester et al., 2023).

Schnack et al. (2016) observed an increase in brain ageing in schizophrenia after the first year of onset, particularly in the subsequent two years, but with a slowdown five years later. Recent findings, however, challenge this proposed relationship between advanced brain ageing and illness duration, especially in samples including more extended periods of illness. Constantinides et al. (2022) did not find any associations between ABA and age of onset or illness duration, suggesting that ABA in schizophrenia may not be primarily driven by disease progression. These findings raise questions about whether the observed gap closes or persists over many years of illness, particularly in medicated patients with established schizophrenia. While a longitudinal study with repeated imaging could provide conclusive answers, as a preliminary step should be assessing groups of patients with first episode or established schizophrenia in an accelerated ageing design.

To date no study has examined the associations between peripheral inflammatory biomarkers, central neurometabolites and advanced brain ageing in schizophrenia. Understanding these potential neurobiological determinants is an important research goal to aid the identification of novel targeted therapies. Theories of progressive brain changes in schizophrenia suggest the involvement of the glutamate system, oxidative stress, and inflammatory pathways, yet the specific contribution of each to ABA have not yet been explored. Magnetic resonance spectroscopy (MRS) measures of glutamate and glutathione (GSH) from the anterior cingulate cortex (ACC) have emerged as reliable indicators of disruptions in glutamatergic signalling and oxidative stress dysfunction in schizophrenia (Egerton et al., 2020, Merritt et al 2021; Das et al., 2019; Dempster et al., 2020; Murray et al., 2023). Additionally, certain inflammatory markers such as interleukin-6 (IL-6) and c-reactive protein (CRP) have demonstrated reliability in indicating the involvement of inflammatory processes in the pathophysiology of schizophrenia and have been linked to brain structural variations within the disorder (Williams et al., 2022; Lalousis et al., 2023).

However, relating model-based estimates of ABA to other mechanistic markers, is challenging and it is important to study the associations based on participant-level contributors for the estimated brain-age. One approach towards extracting participant level features that contribute to a model is to employ the SHapley Additive exPlanations (SHAP) approach (Lundberg & Lee, 2017; Ballester et al., 2023) and measure the marginal contribution of each structural feature to a given prediction. This approach allows the identification of key contributors within patient groups and relate them to other biological markers of interest, providing insights into the potential mechanisms influencing ABA in patients.

In this study, our primary objectives are to investigate whether (1) the brain age gap in schizophrenia varies with the stage of illness (first episode vs. established schizophrenia) when compared to normal brain ageing and (2) whether cortical glutamatergic and antioxidant markers, along with peripheral inflammatory markers, are related to the degree of advanced brain-ageing in schizophrenia. No previous studies relate MRS and inflammatory markers to ABA in schizophrenia and we propose that in identifying potential mechanistic contributors, we lay the groundwork targeted interventions that promote brain health in individuals with schizophrenia.

## 2 Methods

### 2.1 Participants

Participants were recruited as part of the “The Study of Psychosis and the Role of Inflammation and GABA/Glutamate” (SPRING) (Deakin et al., 2024). In total, 207 participants were recruited across three sites: Cardiff, Manchester, and Nottingham. The study included 58 first episode psychosis (FEP) patients who were minimally medicated (<12 weeks), aged 18-41, and within 5 years of symptom onset, 76 established psychosis patients (EST) with symptoms present for more than 10 years and aged 27-55. 73 healthy controls (HC) were age and sex matched to the participants. For this analysis, only participants recruited to the Cardiff and Manchester sites were included due to variation in magnet strength (3T GE/Siemens in Cardiff and Manchester, 7T Phillips in Nottingham). Participants were excluded from the analysis if they did not have a combination of blood-based inflammatory markers, 1H-MRS, and 3T structural MRI at baseline with an image quality rating >75% (assessed by CAT-12 toolbox version r1830; http://dbm.neuro.uni.jena.de/cat12/). The resulting sample consisted of 112 participants, including 34 FEP (10 Female), 36 EST (10 Female), and 41 HC (10 Female) (Table 1). Additional measures for patients’ current antipsychotic usage were collected, including defined daily dose, and an 11- point rating of lifetime antipsychotic exposure measure reflecting both dose and duration of medication with fixed criteria for each level.

**Table 5.1:** Factor loadings for each component.

|  | PC1 | PC2 | PC3 | PC4 | PC5 |
| --- | --- | --- | --- | --- | --- |
| **IFN-γ** | **0.36** | 0.35 | -0.33 | 0.08 | 0.27 |
| **IL-10** | 0.27 | -0.15 | **0.49** | -0.30 | 0.39 |
| **IL-12** | 0.04 | 0.07 | -0.06 | 0.42 | **0.45** |
| **IL-13** | 0.07 | **0.41** | 0.24 | 0.32 | -0.20 |
| **IL-1B** | 0.05 | 0.44 | **-0.39** | -0.22 | 0.36 |
| **IL-2** | 0.16 | 0.37 | 0.29 | **0.43** | -0.13 |
| **IL-4** | 0.04 | **0.33** | -0.14 | **-0.54** | -0.22 |
| **IL-6** | **0.56** | -0.21 | -0.12 | 0.04 | -0.18 |
| **IL-8** | 0.13 | **0.38** | 0.36 | -0.25 | **-0.29** |
| **TNF-a** | **0.47** | **-0.11** | 0.29 | -0.14 | 0.26 |
| **CRP** | **0.46** | **-0.21** | -0.34 | 0.10 | -0.38 |

PC1: Pro-inflammatory Factor
Key Features: ↑ IL-6, ↑ TNF-a, ↑ CRP
PC2: Anti-inflammatory Factor
Key Features: ↑ IL-13, ↑ IL-4, ↓ CRP, ↓ IL-6
PC3: Immuno-modulation Factor
Key Features: ↑ IL-10, ↓ IL-1B
PC4: T-Cell Modulation Factor
Key Features: ↑ IL-2, ↓ IL-4
PC5: Th1 Immune Response Factor
Key Features: ↑ IL-12, ↓ IL-8

**Table 5.2:**
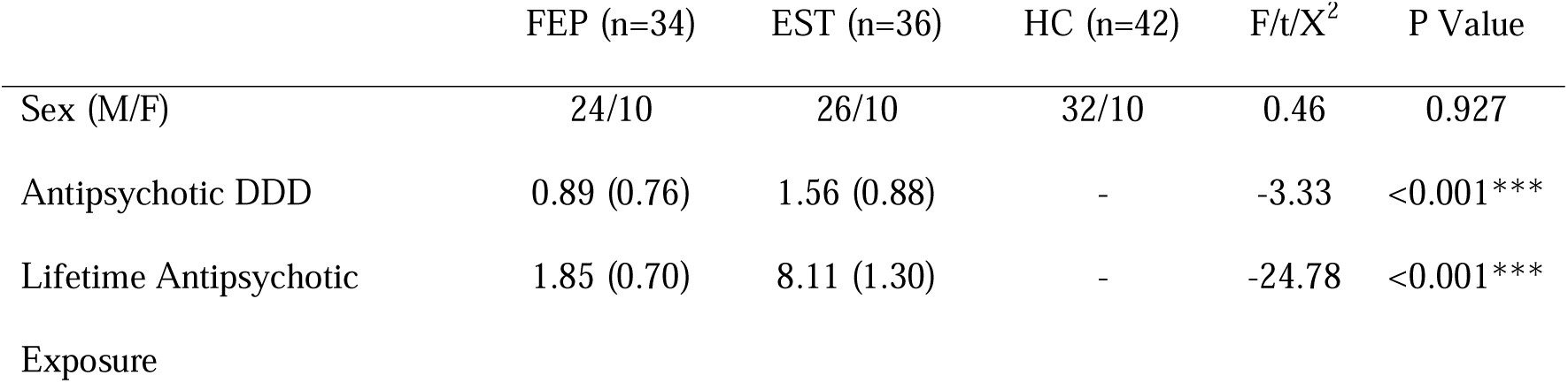
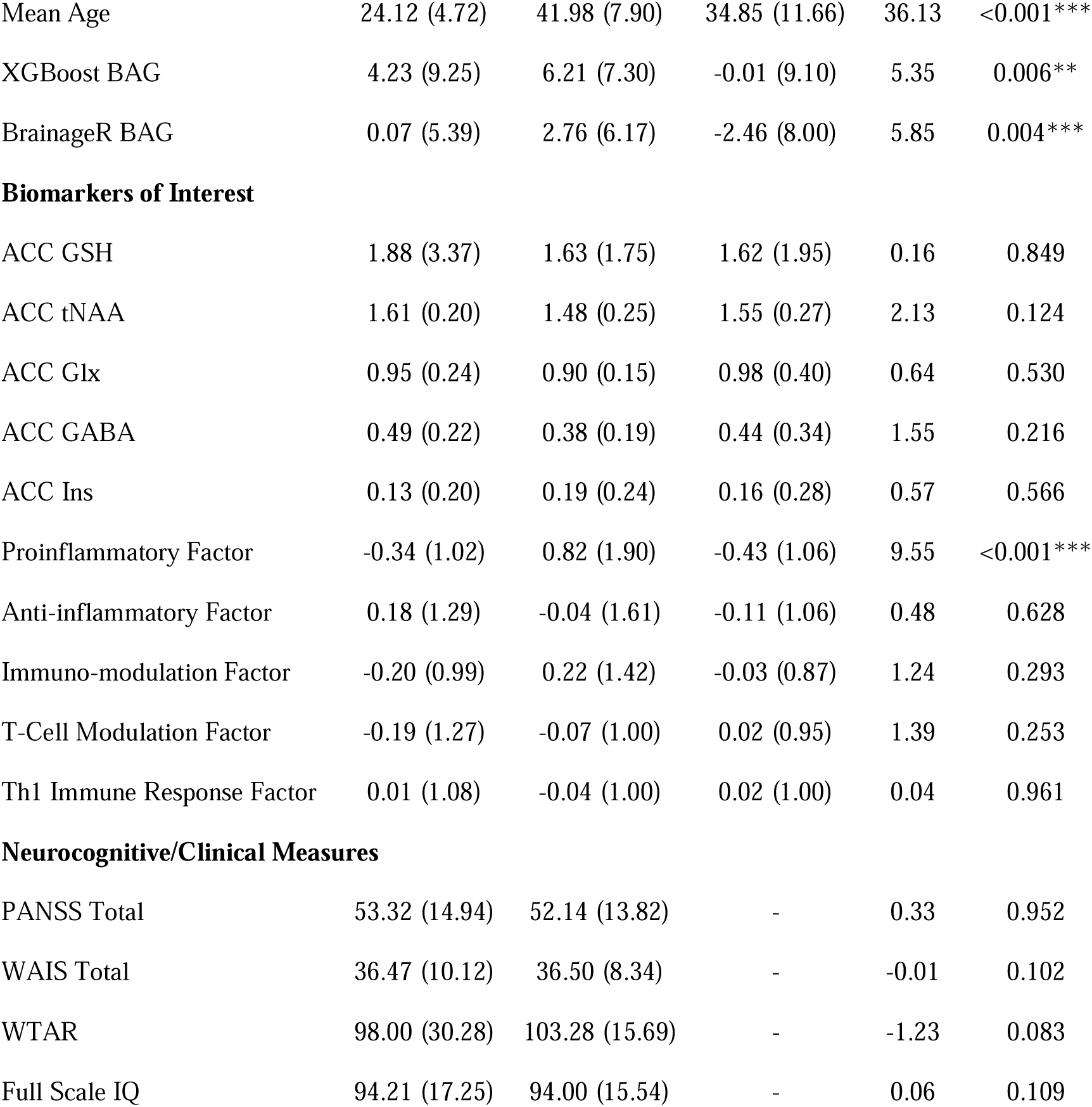
Demographic Information and Markers of Interest in participant groups.

### 2.2 Data Acquisition

Participant symptoms were assessed using the Positive and Negative Syndrome Scale (PANSS) (Kay et al., 1987). Scores for 5 PANSS dimensions: the positive dimension, negative dimension, anxio-depressive dimension, excitement dimension and disorganisation/other dimension were all calculated via summing of relevant symptom scores (Yazaji et al., 2002). Neurocognitive function was evaluated using the Wechsler Adult Intelligence Scale III for patients with schizophrenia, which included subtests such as “Information,” “Arithmetic,” “Block Design,” and “Digit Symbol Test” for processing speed. Premorbid IQ was assessed using the Wechsler Test of Adult Reading (WTAR) (Wechsler, 2001).

Venous blood was collected and centrifuged within one hour at 1300–2000g for 10 min. In total 11 inflammatory markers were collected – IL-1B, IL-2, IL-4, IL-6, IL-8, IL-10, IL-12, CRP, IFN-y and TNF-a. Cytokines were measured in duplicate using Meso Scale Discovery (MSD) V-plex immunoassays, Pro-inflammatory Panel 1 (human) kit (MSD, Maryland, USA), and MSD Vascular Injury Panel 2 human kit weas used to measure CRP levels.

T1-weighted structural neuroimaging was conducted using a standard acquisition protocol. The Cardiff site collected data with a 3T GE MRI scanner, and the Manchester site collected data on a 3T Philips MRI scanner. Further 1H-MRS neuro-metabolite data was acquired at each site following a standardised sequence (see supplement for details). A single voxel was placed in the bilateral dorsal anterior cingulate cortex (dACC) superior to the genu of the corpus callosum (supplementary figure s1). A point resolved spectroscopy (PRESS) 1H-MRS sequence (TE/TR = 35/2000 ms, 2000 Hz bandwidth, number of points = 1024, number of averages = 128, Voxel size = 35 x 40 x 20 mm^3^) was used to acquire water-suppressed spectra as well as a water-unsuppressed spectrum for quantification. Additional Mescher-Garwood (MEGA)-edited PRESS data for GSH were collected from the same voxel (TE/TR = 130/2000 ms, 2000 Hz bandwidth, number of points = 1024 (Manchester), 5000 Hz bandwidth, number of points = 4096 (Cardiff), number of averages = 256).

### 2.3 Pre-processing and Site Correction

Raw T1 structural MRI data was assessed for data quality using the open-source CAT- 12 toolbox (version r1830; http://dbm.neuro.uni.jena.de/cat12/), and participants with a raw image quality rating <75% were excluded from further analysis (see supplementary methods). Images were pre-processed and segmented using the *recon-all* command in Freesurfer 7.2.0. Relevant features were extracted using *asegstats2table* within Freesurfer and the HCP multimodal atlas (Glasser et al., 2019). In total, 1,118 features were extracted, covering volume, area, and thickness measurements.

PRESS and MEGA-PRESS data were pre-processed and quantified using the Spectroscopy Analysis Tools (SPANT) software package in R (https://github.com/martin3141/spant; Wilson 2021a). SPANT pre-processing included coil combination, identification of bad averages, and frequency and phase correction (Wilson, 2018). Data were quantified using ABFit model fitting (Wilson 2020), and basis sets were simulated within SPANT for the metabolites: N-acetylaspartate (NAA), N-acetylaspartylglutamate (NAAG), glutamate, glutamine, creatine, phosphocreatine, glycerophosphocholine, phosphocholine, myo-inositol (Ins), scyllo-inositol, GABA, aspartame, taurine, glucose, glutathione (GSH), and glycine. Some metabolite concentrations were then combined to estimate “total” (t) concentrations: tNAA (NAA+NAAG), tCr (creatine + phosphocreatine), Glx (glutamate + glutamine). Of the metabolites measured, 5 were selected for further analysis: GSH (derived from MEGA-PRESS edited MRS), Glx, tNAA, GABA and Ins (derived from PRESS MRS). These were selected due to their high data quality (Cramer Rao Lower Bounds <20% in most participants) and their relevance to schizophrenia.

ComBat harmonization was employed to correct for MRI scanner variances, batch effects, and site effects. ComBat was run separately for the T1 structural, MRS, and blood-based biomarkers data. Correction was implemented using the neuroCombat package (https://github.com/Jfortin1/neuroCombat_Rpackage). Covariates of age, sex, and disease status were protected during the removal of site/scanner effects. Empirical Bayes was set to false for the MRS and blood-based biomarkers due to the number of features being substantially smaller than the number of participants.

Principal component analysis was conducted in order to reduce the dimensionality of the inflammatory marker data. The kaiser criterion was employed to select the optimal number of components while explaining the greatest amount of variance in the data.

### 2.4 Brain Age Prediction

The publicly available “brain-age” prediction model developed by Kaufmann et al. (2019) was used to assess advanced brain ageing in our dataset. Pre-trained gradient-boosted trees generated by the XGBoost method, provided by the authors, are available online (https://github.com/tobias-kaufmann/brainage). Full details of the model training can be found in (Kaufmann et al., 2019), however in brief, two separate models were trained (one for males and one for females) on a total of 35,474 individuals aged 3-89 years. The average chronological ages from the pretrained models were 48.01 for males and 46.63 for females. Model performance was assessed by the mean absolute error (MAE) between predicted brain age and chronological age and the Pearson correlation coefficients (r) between predicted brain age and chronological age.

The resulting XGBoost models were compared to the performance of a separate pretrained model, BrainAgeR (https://github.com/james-cole/brainageR), to ensure prediction accuracy. BrainAgeR employs Gaussian Process regression to estimate brain age using raw T1 scans in nifti format, potentially reducing the confounding effects of atypical brain morphology and removing variance introduced by differences in pre-processing (Cole et al., 2017). The original BrainAgeR model was trained on 3377 healthy individuals aged 18-92 (mean age = 40.6 years).

As the accuracy of predicted brain age varies with sample age, an age-correction procedure proposed by Beheshti et al. (2019) was employed. This procedure fits a linear model between predicted age and age of healthy participants to adjust for age-related confounds. Then, an age-corrected predicted age is derived for both HC and SCZ samples based on the slope, intercept, and predicted age extracted from the HC group. This method ensures that the model has a consistent level of error across the whole age range of healthy participants which can then be applied to the schizophrenia sample. This corrected prediction was used to compare brain age gap BAG across the groups. Subsequent analyses of neurometabolic and inflammatory marker associations were conducted using the raw, uncorrected, brain-age predictions generated from the XGBoost model with age and age^2^ included as covariates in the models to reduce the likelihood of over-correction.

### 2.5 Shapley’s Additive Explanations (SHAP)

To better understand the contribution and importance of individual structural brain measures for making brain age predictions, Shapley’s Additive Explanations (SHAP) method was used. SHAP is a method based on game theory that extracts marginal contributions of features from predictions. The SHAP value for a specific feature for an individual prediction represents the difference in the prediction when that feature is left out of the decision tree (marginal contribution). The SHAP for XGBoost package in R (https://cran.r-project.org/web/packages/SHAPforxgboost/index.html) was used in this case (Lundberg and Lee, 2017; Ballester et al., 2023). SHAP values represent the contribution of each feature to the deviation of the prediction from the mean age of the database (the starting point of the XGBoost model).

### 2.6 Statistical Analysis

Brain age gap (BAG) was calculated as brain-predicted age minus chronological age. This was computed for each SCZ participant and used as the outcome variable. Although sex-specific prediction models were built, the generated BAG values were pooled across males and females for subsequent statistical analyses within each cohort. This was done to improve statistical power as previous work has shown similar correlations with chronological age in each sex (Bacas et al., 2023). BAG was assessed to identify significant brain age differences between the two stages of schizophrenia (FEP and EST) and age and sex-matched healthy controls.

The EST and FEP schizophrenia groups were then combined into one cohort to improve statistical power in analysis of the relationship with biomarkers of interest. Partial correlation matrices were then generated for this combined schizophrenia cohort and healthy controls to assess the associations between uncorrected BAG, MRS-derived neurometabolites, blood-based biomarkers of inflammation, and clinical/neurocognitive characteristics. In each matrix, both age and age-squared were included as covariates to correct for the systemic age-related bias in brain age prediction. Similar matrices were generated for the top 10 performing SHAP features in each model to assess the association between these features of interest and brain-derived neurometabolites or blood-based inflammatory markers. This method can help delineate the underlying mechanisms of association between these features (supplementary results).

## 3 Results

### 3.1 PCA Dimensionality Reduction

Principal component analysis of all inflammatory markers was conducted to reduce the dimensionality of the dataset. Based on the Kaiser criterion (eigenvalues >1), five principal components were retained for further analysis. These five principal components capture approximately 67% of the total variance.

### 3.2 Model Comparisons

Overall BrainageR marginally outperformed the XGBoost model on typical measures of model performance. It had a lower mean absolute error (6.75 years vs 7.17 years), but R^2^ association between brain predicted age and chronological age was the same for both models. As such further analysis was conducted on the XGBoost model outputs as Shapley’s additive explanations is more informative with gradient-boosted tree models.

### 3.3 Brain-Age Predictions

Overall, using the XGBoost model predictions, brain-ageing was seen to be accelerated in the EST group compared to HC (*p* = 0.005), but the FEP group was not significantly different from either the EST or HC groups (*p* = 0.605, *p* = 0.088 respectively). The same result was found for the BrainageR model (Figure 2).

**Figure 1:**
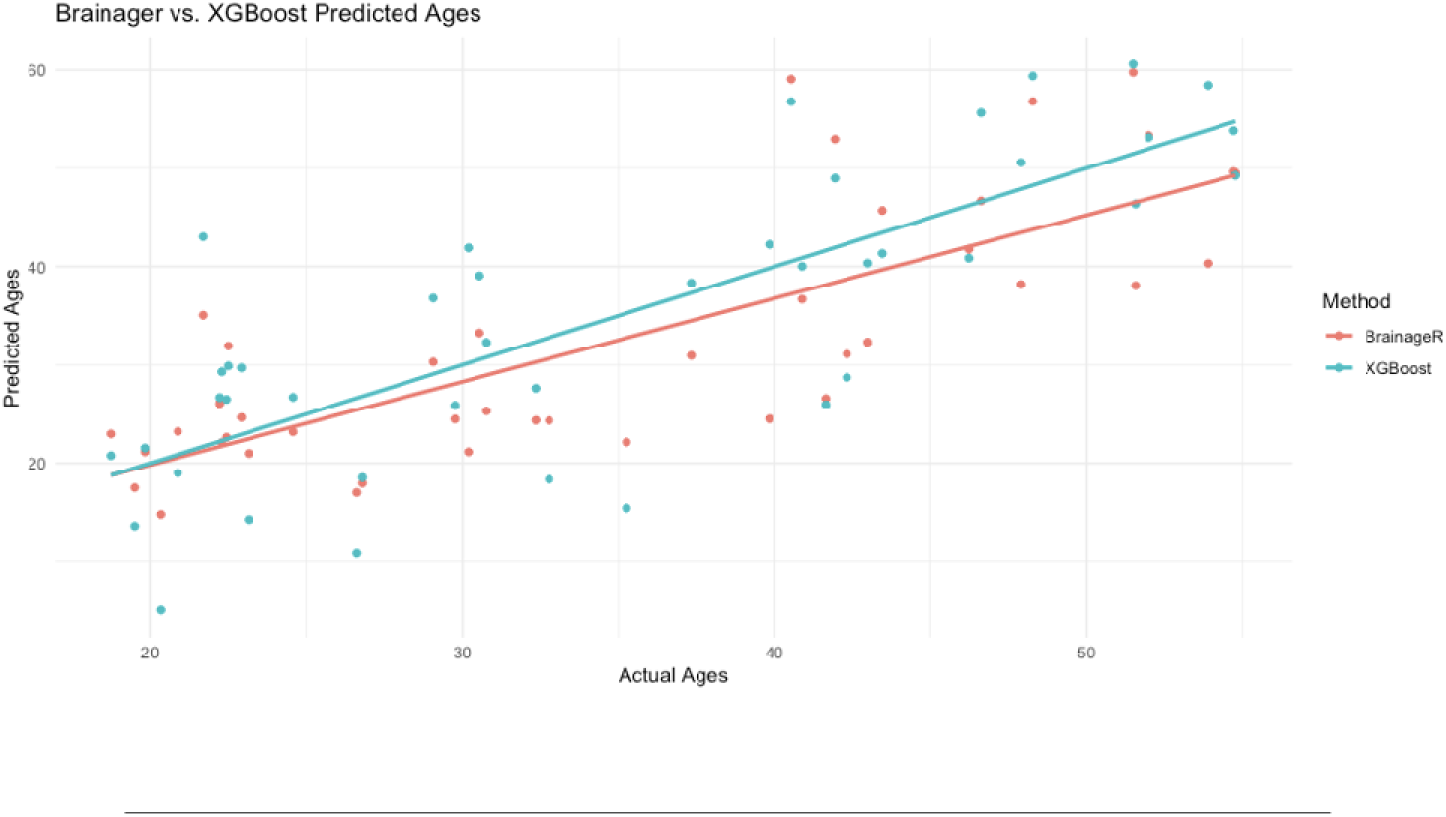
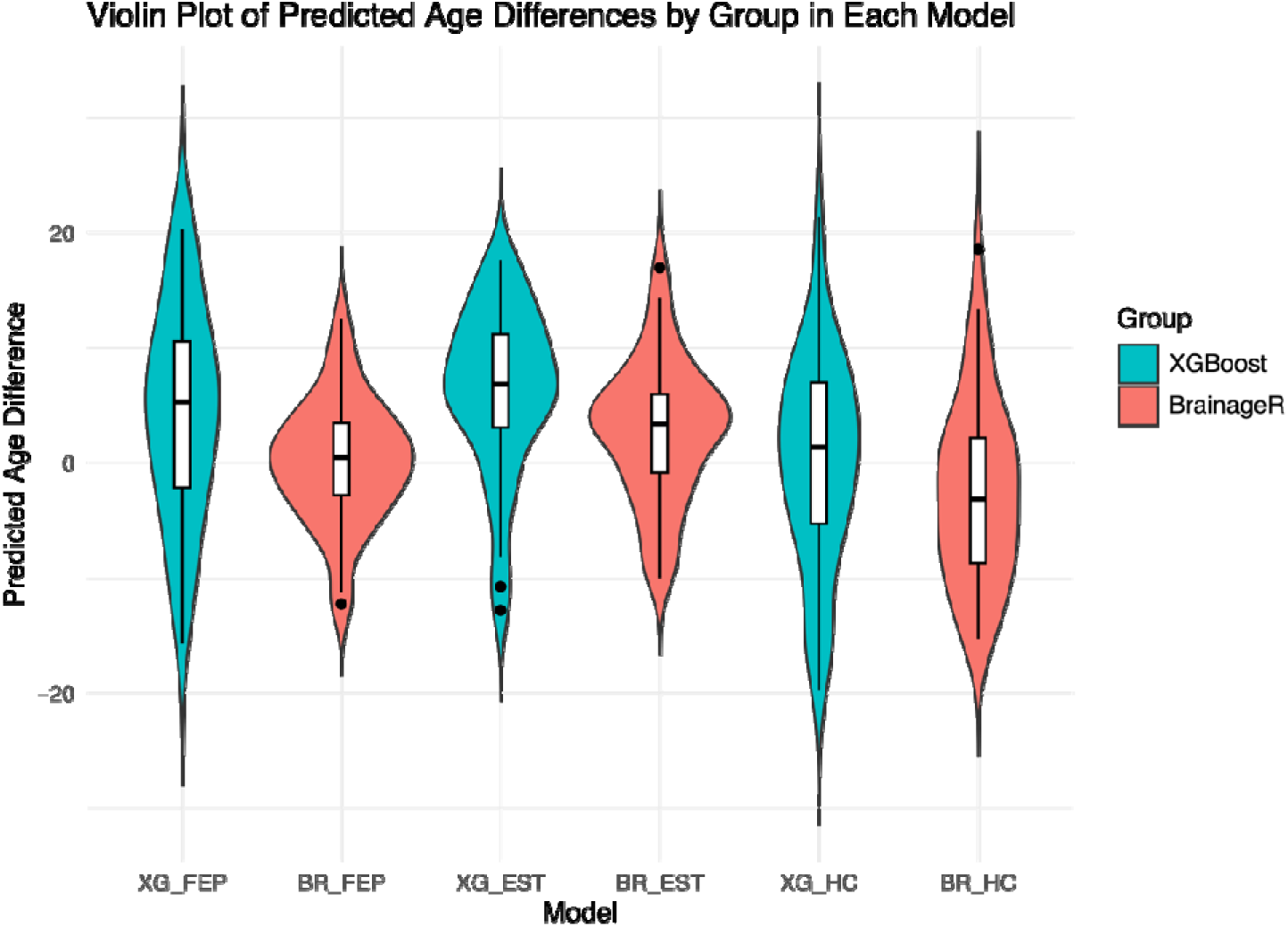
Performance of XGBoost and BrainageR Models. Top: Comparison of model performances: chronological age vs model predicted age of HCs. BrainageR R^2^ = 0.62; XGBoost R^2^ = 0.62 Bottom: Violin plots displaying predicted age difference (brain predicted age – chronological age) for each group in each model. Mean absolute error for healthy controls BrainageR = 6.75 years; XGBoost = 7.17 years

**Figure 2:**
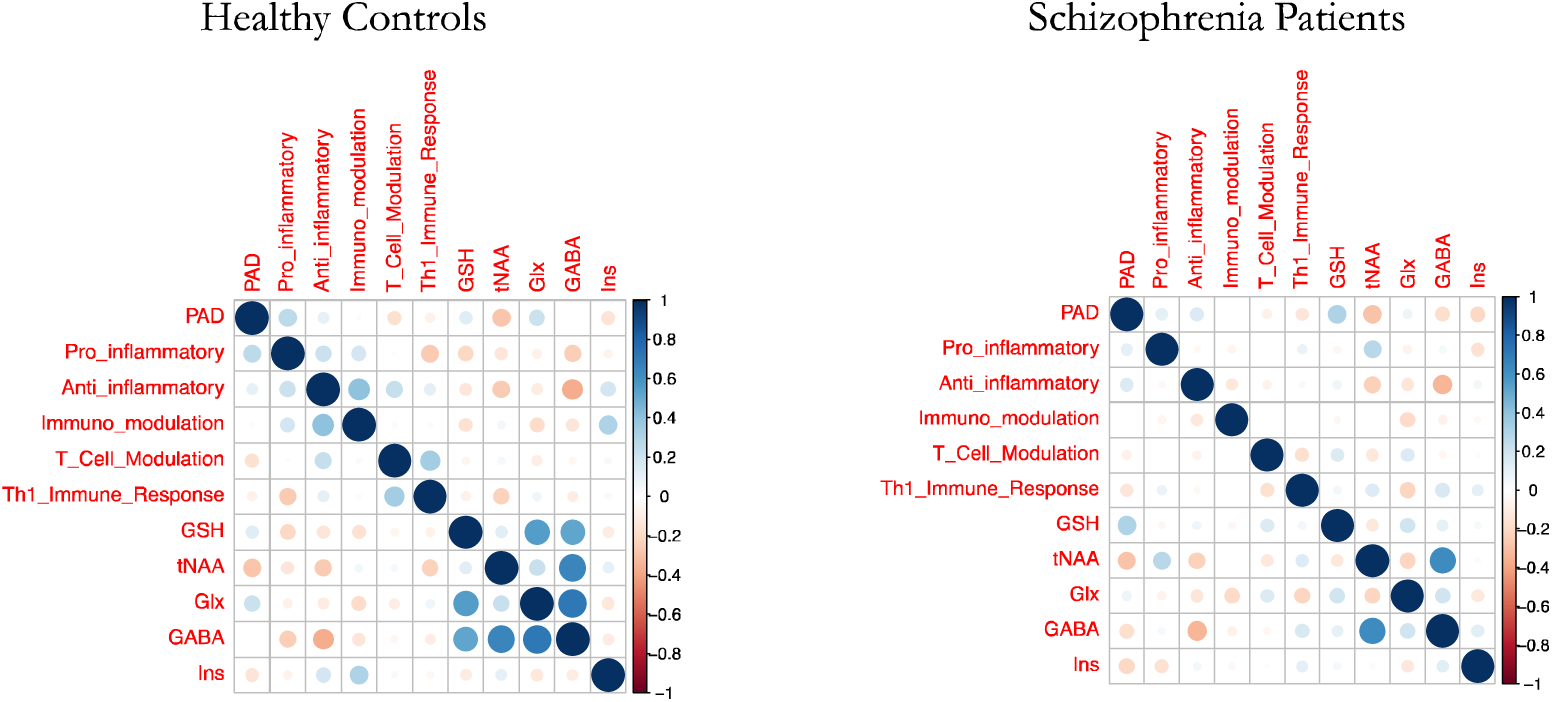
Correlation Matrices Demonstrating Associations Between BAG and Biomarkers of Interest. Blue dot indicates positive correlation, orange indicates negative. Diagonal displaying self- correlation is darkened out. Dot size represents strength of correlation.

**Figure 3:**
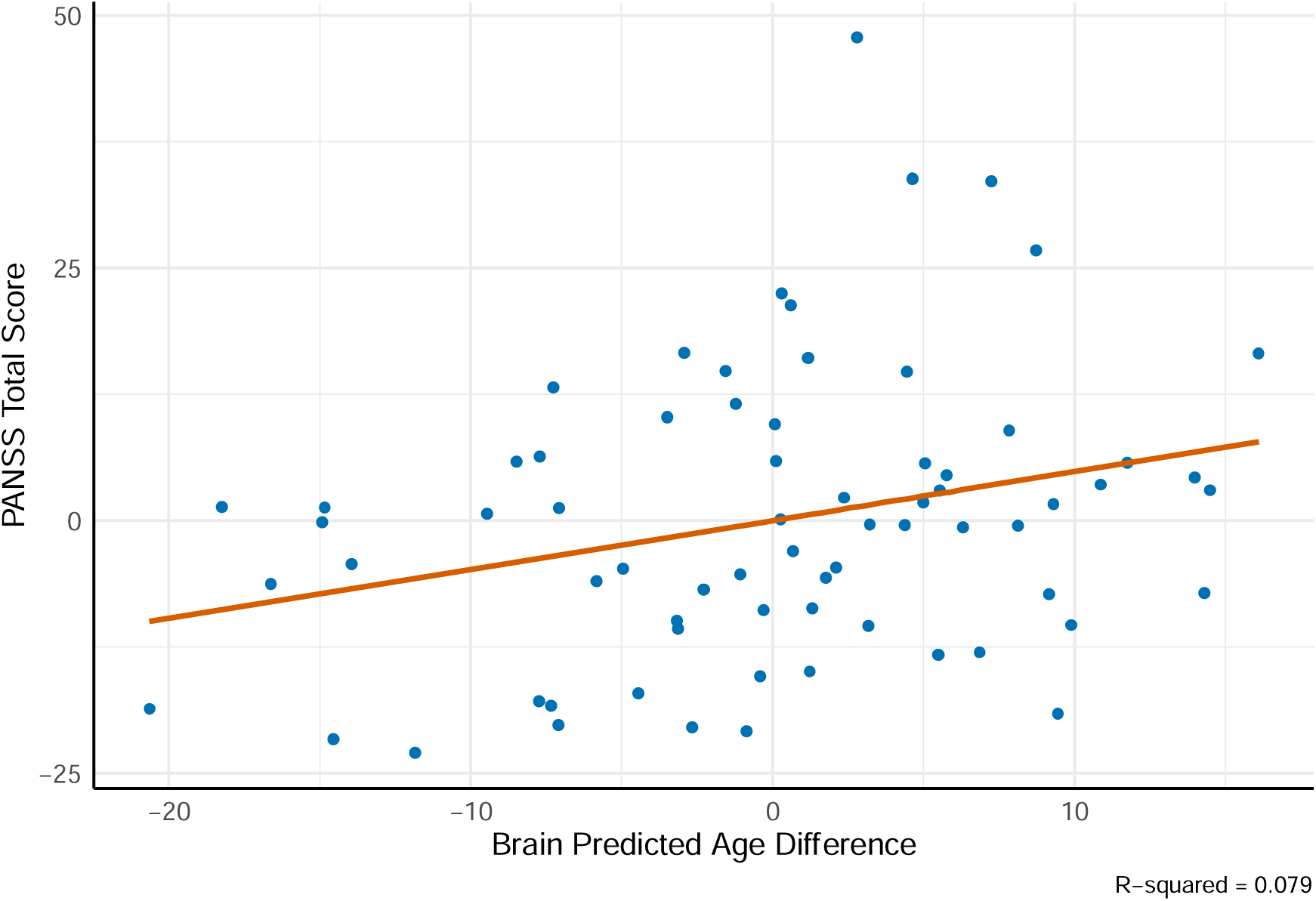
Relationship Between BAG and PANSS Total Scores.

**Figure 4:**
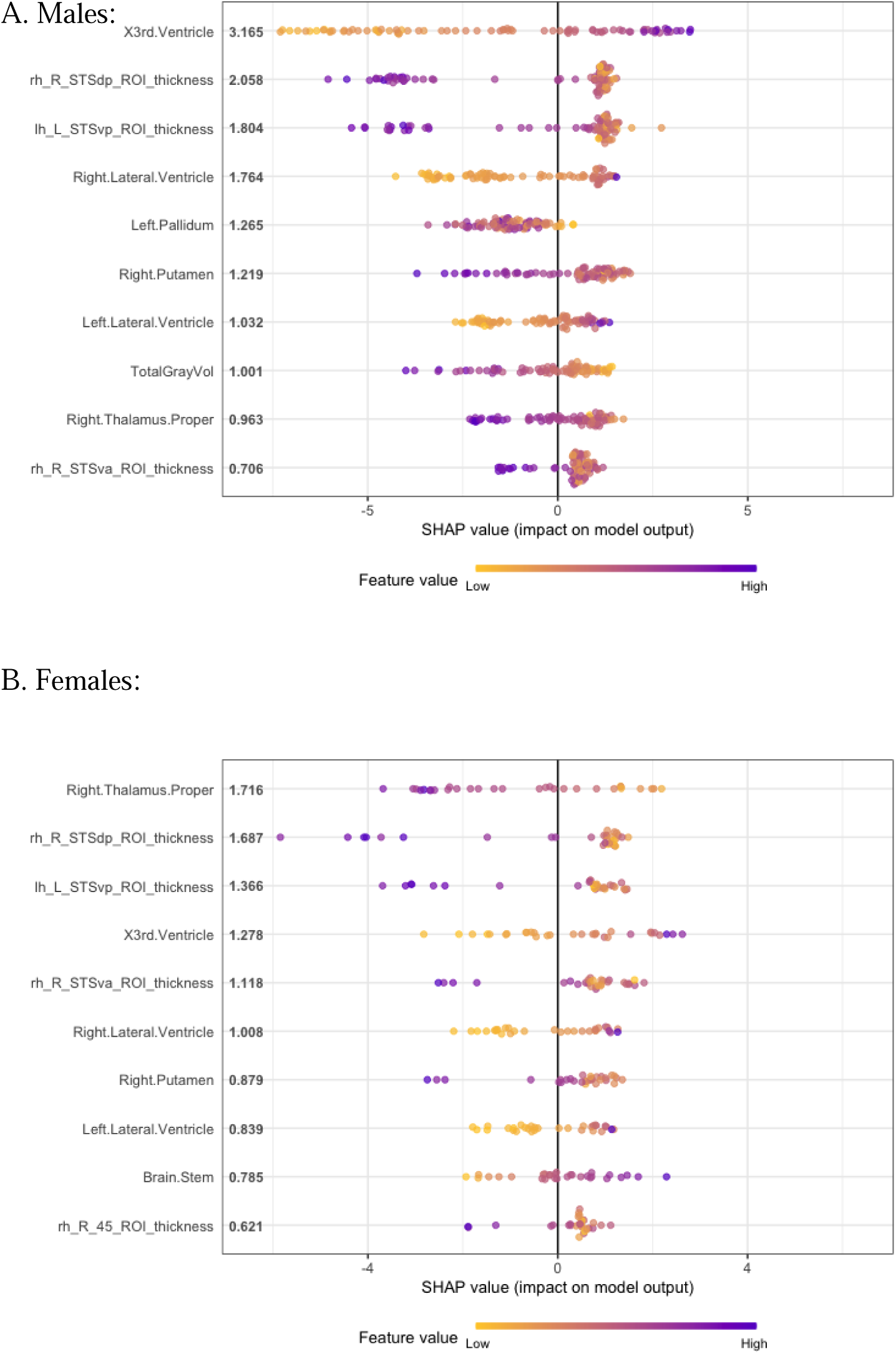
Top 10 SHAP features for each model. *Rh* Right hemisphere; *lh* left hemisphere; *STS* superior temporal sulcus; *dp* dorsal posterior; *vp* ventral posterior; *va* ventral anterior; *ROI* region of interest.

### 3.4 Biomarker Associations with Brain age Gap (BAG)

Partial Pearson correlation matrices, controlling for the effects of age and age^2^, were generated for each group to assess associations between BAG difference and biomarkers. Within the combined group the biomarkers presenting with the greatest correlation with predicted age difference were total N-acetylaspartate (r(66) = -0.287, *p* = 0.018) and GSH (r(66) = 0.301, *p* = 0.013). In contrast, the HC group demonstrated no significant associations with any biomarker of interest. No blood-based biomarker factor was significantly correlated with BAG in either SCZ or HC, however when looking at the whole dataset (SCZ+HC), the pro-inflammatory factor was significantly positively associated with BAG (r(108) = 0.227, *p* = 0.017) (see supplementary results). Furthermore, predicted-age difference was not associated with defined daily dose of antipsychotic medication (r(68) = 0.034, *p* = 0.786) nor lifetime anti-psychotic exposure (r(66) = -0.004, *p* = 0.976).

### 3.5 Neurocognitive Associations with Brain Age Gap

Neurocognitive scores, including PANSS subscales, WAIS scores, IQ and WTAR, were included in a partial correlation matrix alongside BAG. The effects of age and age^2^ were controlled for. Overall, no significant associations were found between BAG and any of the cognitive scale scores within the combined schizophrenia group. However, significant positive associations were found with PANSS general (r(66) =0.299, *p* = 0.013), total (r(66) = 0.281, *p* = 0.020) and anxio-depressive scores (r(66) = 0.238, *p* = 0.050).

### 3.6 Shapley’s Additive Explanations

To better understand the contribution of individual features to brain-predicted age, irrespective of diagnosis, SHAP values were calculated for each of the 1,118 features and averaged across all participants, regardless of group. The top ten most relevant features based on mean absolute contribution to the prediction were extracted from the male and female models. In total 12 unique features were identified (9/10 features overlapped between models). One-way ANCOVA, with sex, age and age^2^ as covariates, was conducted to assess how these features differed between groups. None of the individual features differed significantly between groups i.e. no individual feature from the top 12 was significantly differently to brain age estimation in the schizophrenia groups than the healthy controls, indicating that the same morphological feature were driving the brain-predicted age measure in both groups.

## 4 Discussion

The present study aimed to identify if advanced brain ageing was evident across different phases of psychosis. Overall, it was found that the established schizophrenia group showed more advanced brain ageing compared to controls, while the first episode psychosis group did not differ significantly from controls. The top features related to advanced brain age, including ventricular enlargement, subcortical thalamic and total grey matter volume showed no significant differences between patients and controls suggesting cumulative minor alterations similar to ‘normal ageing’ may contribute to advanced brain ageing in psychosis. In the combined psychosis group, more advanced brain ageing was associated with more severe symptoms, increased glutathione, and decreased N-acetyl aspartate but did not show evidence of a relationship with peripheral markers of inflammation, notably pro-inflammatory markers.

Our findings are consistent with prior research findings (Koutsouleris et al., 2015; Ballester et al., 2021) however challenge some studies that focused on the early phases of psychosis, which reported an average advancement in brain ageing by 2-3 years (Schnack et al., 2016; Hajek et al., 2017; Kolenic et al., 2018). Our results do indicate a trend towards a higher brain-age gap in the FEP group compared to HC, indicating that the effect size of accelerated brain ageing in FEP is smaller than in established cases, thus requiring a larger sample size for it to be detected. In fact, the absence of a significant difference in brain-age gap between first episode and established patients also supports the presence of subtle but small effect of ABA in FEP in a sample of this size. One reason for this small effect could be the presence of more neurobiologically heterogeneous subgroups in the FEP sample, compared to the established cases.

Unlike previous work identifying total grey matter volume as the primary underlying feature driving increased BAG in schizophrenia (Ballester et al., 2023). We did not find any individual SHAP feature to differ between schizophrenia and controls. This may indicate that, at least in the dataset presented, the cumulative impact of subtle brain alterations contributes to the advanced brain ageing seen in schizophrenia. Indeed, many studies have implicated a number of widespread variations in brain volume, area, and thickness in schizophrenia. Madre et al., 2020 identified extensive cortical volume and thickness reductions across the whole brain in schizophrenia, alongside surface area reductions in the superior frontal cortex. Similarly, in their study of 4,474 individuals with schizophrenia, the ENIGMA consortium identified cortical thinning in all Desikan-Killiany (DK) atlas regions bar the bilateral pericalcarine region and smaller cortical area in all DK atlas regions bar the bilateral isthmus cingulate region (Van Erp et al., 2018).

To our knowledge this is the first study to relate peripheral inflammatory markers and MRS-derived neurometabolites to advanced brain ageing. We identified positive associations with ACC GSH and negative associations with ACC tNAA in the combined-schizophrenia group. GSH is the brain’s dominant antioxidant, measurement of which is the primary method used to assess oxidative stress within the brain (Palaniyappan et al., 2021). Past research has suggested that there is a reduction in brain GSH within psychosis and therefore, people with schizophrenia are more vulnerable to the damaging effects of free radicals (Do et al., 2000; Kumar et al., 2020). In our recent meta-analysis, we identified significant GSH reductions in the ACC of individuals with established schizophrenia but not first episode, indicating potential dysregulation in oxidative stress pathways in the later stages of the disorder. Though we did find a significant association with study year – when older studies, likely employing outdated methodologies, were removed, GSH levels in the established group did not differ from controls (Murray et al., 2024).

The positive association of GSH with brain age gap identified here may be counter to some previous findings. Notably, certain studies have reported GSH increases in individuals experiencing first episode psychosis (Limongi et al., 2021; MacKinley et al., 2022). The rise in GSH has been explained as a compensatory reaction to heightened oxidative stress within the brain. Consequently, individuals with schizophrenia in whom processes that promote accelerated brain ageing are active (e.g. neurotoxic oxidative stress), a trigger for a compensatory rise in GSH may operate. Given the cross-sectional nature of our study with two stage specific groups, we are not able to conclude if the elevated GSH levels indeed confer any clinical or functional advantage in patients, though prior studies (Dempster et al., 2020; Mackinley et al., 2022) provide some support to this notion.

N-acetyl-aspartate serves as a commonly utilized indicator of neuronal health (Rae, 2014). Numerous studies have reported diminished levels of tNAA in conjunction with neuronal loss or impaired neuronal metabolism (Hardy et al., 2011; Malaspina et al., 2016; DeMayo et al., 2023). As such, the observed decrease in tNAA linked to the elevated predicted-age difference may implicate ongoing neuronal degradation. Additionally, tNAA is known to exhibit a substantial decline with advancing age. Even though age was controlled for in all our analyses, it remains plausible that this reduction may still mirror the impacts of ageing on brain-predicted age difference.

We did not identify any association between BAG and peripherally measured inflammatory markers in either cohort. Suggesting that peripheral inflammation may not be associated with advanced brain ageing. To date several studies have identified associations between brain structure and blood-based inflammatory markers though findings have been disparate. In one study, patients presenting with “classic inflammation” demonstrated widespread grey matter volume reductions in a number of brain regions including the anterior cingulate and frontal gyri (Lalousis et al., 2023), alternatively grey matter thickening was seen in patients within a high “inflammatory proband” compared to those in the low inflammatory group (Lizano et al., 2021). Further studies have found no association between peripheral inflammatory markers and brain structure but did find an association with monocyte gene expression and cortical thickness (Cui et al., 2023). These results bring into question the validity of using blood-based inflammatory markers to identify structural and functional differences in the brain and highlights the need for improved methods for non-invasively assessing neuro-inflammation.

The positive associations between BAG and PANSS total, general, and anxiodepressive subscales mirror findings from previous works, with a recent meta-analysis highlighting PANSS scores on all subscales to be increased with advanced brain ageing (Blake et al., 2023), thus highlighting the clinical relevance of this phenomena. Similar to previous studies (Koutsouleris et al., 2014; Kolenic et al., 2018; Wang et al., 2021), we found no association with defined daily dose of antipsychotic medication also suggests that this advancement in brain-age is likely not a result of medicinal intervention. Taken together this highlights the utility of BAG as a neurobiological marker reflecting the severity of psychiatric symptoms in schizophrenia. Clinically targeting advanced brain ageing may offer a novel avenue for interventions aimed at improving overall brain health and mitigating symptom burden in individuals with schizophrenia.

While this study provides valuable insights, certain limitations are worth consideration. The cross-sectional nature of the study limits our ability to establish causal relationships. Longitudinal studies with extended follow-up periods are crucial to elucidate the trajectory of advanced brain ageing over the course of schizophrenia. Furthermore, our sample size may be underpowered to detect group-specific associations between biomarkers and BAG. Additionally, the study’s focus on neurometabolites in the anterior cingulate cortex calls for future investigations into other brain regions to comprehensively understand the neurobiological basis of accelerated ageing in schizophrenia.

In conclusion, advances our understanding of advanced brain ageing across different stages of psychosis, suggesting more pronounced advancements in established schizophrenia stages compared to first episode psychosis. Our findings suggest that the cumulative impact of subtle brain alterations, rather than distinct feature changes, contribute to the observed acceleration. Associations with neurometabolites, particularly GSH and tNAA, highlight the complex interplay between oxidative stress and overall brain health within psychosis. The lack of significant links between advanced brain ageing and peripheral inflammatory markers highlights the need for further research to explore alternative pathways of neuroinflammation. The correlations between advanced brain ageing and symptom severity emphasise the potential of brain age as a marker for clinical interventions aimed at improving brain health and alleviating symptoms in schizophrenia. Future longitudinal studies with larger sample sizes and broader analysis of specific brain regions are essential to progress our understanding of the trajectory and underlying mechanisms of brain ageing in psychosis.

## 7 Author Contributions

AM conceived the initial research project and collated existing data. Data collection and processing of inflammatory markers were conducted by the SPRING team (BD, PFL, MZK, and SW) prior to analysis. AM conducted all statistical analyses, research, and drafted the article. PB assisted AM in producing SHAP values, while MW aided in extracting MRS data. All authors critically revised the article, providing support on content and structure.

## Data Availability

All data produced in the present study are available upon reasonable request to the authors

