## Supplement for "Probing the Biological Underpinnings of Advanced Brain Ageing in Schizophrenia" (from another version of this preprint: version 1, 2024-07-05)

**Supplementary Methods:**

Data Acquisition:

All data for each participant was acquired in one day, with a morning of cognitive tests and interviews, then blood acquisition, followed by an afternoon of imaging procedures.

sTable 1: Scanners and Coils

| Site | Scanner | Field | Receive Coil |
| --- | --- | --- | --- |
| Cardiff | GE HDx | 3.0 T | 8 channel head coil |
| Manchester | Philips Achieva | 3.0 T | 32 channel head coil |

sTable 2: Sequence Parameters and Acquisition Details

| MRS Sequence | TE/TR (ms) | Navg | BW (Hz) | Pts |
| --- | --- | --- | --- | --- |
| PRESS | 35/2000 | 128 | 2000 | 1024 |
| MEGA-PRESS | 130/2000 | 256 | 5000 (Cardiff)  2000 (Manchester) | 4096 (Cardiff)  1024 (Manchester) |

sTable 3: GSH MEGA-PRESS Acquisition

| TE (ms) | Frequency offsets (edit/control) | Edit pulse shape | Length/BW |
| --- | --- | --- | --- |
| 130 | 4.56/1.44 ppm | gauss | 14ms/106Hz |

sTable 4: T1 Anatomical Images

| Site | Sequence | TI/TE/TR (ms) | Matrix | Voxel Size |
| --- | --- | --- | --- | --- |
| Cardiff | FSPGR | 450/88/3.5 | 256x256x192 | 1mm isotropic |
| Manchester | IR-TFE | 1150/3.9/8.4 | 256x256x90 | 0.9x0.9x1.9 mm |

FSPGR: 3D Fast Spoiled Gradient-Recalled acquisition

IR-TFE: Inversion Recovery prepared Turbo (rapid) Field Echo (MPRAGE (7))

sFigure 1: MRS Bilateral ACC Voxel Placement.

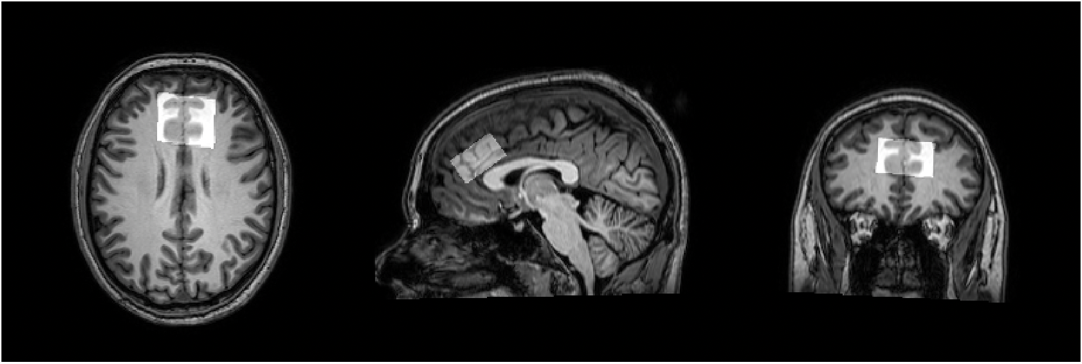

Image Quality Rating:

After segmentation cat-12 will automatically output image quality ratings for each T1 file. The image quality measures describe the properties of the image before CAT12 processing. They were estimated by using the tissue segmentation and were (nearly) independent of subject sex and health status.

sFigure 2: CAT12 Image Quality Ratings

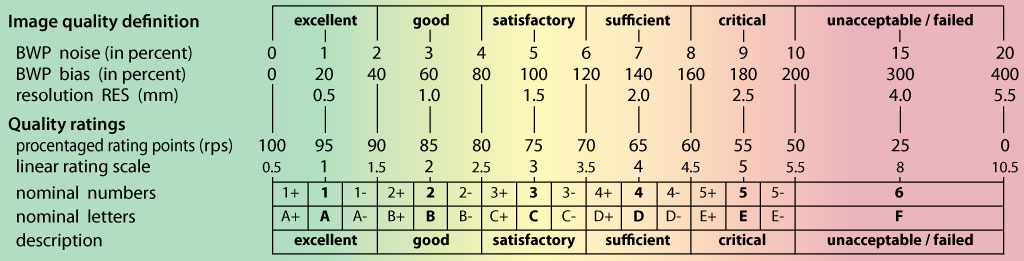

**Supplementary Results:**

sFigure 3: Relationship Between BAG and Defined Daily Dose

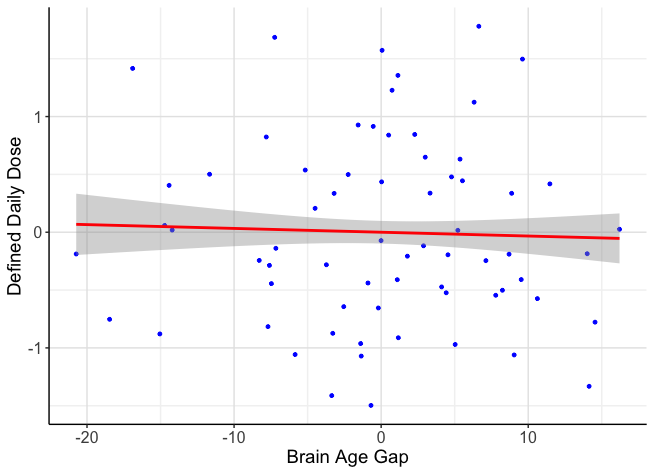

sFigure 4: Relationship Between BAG and Cumulative Antipsychotic Score

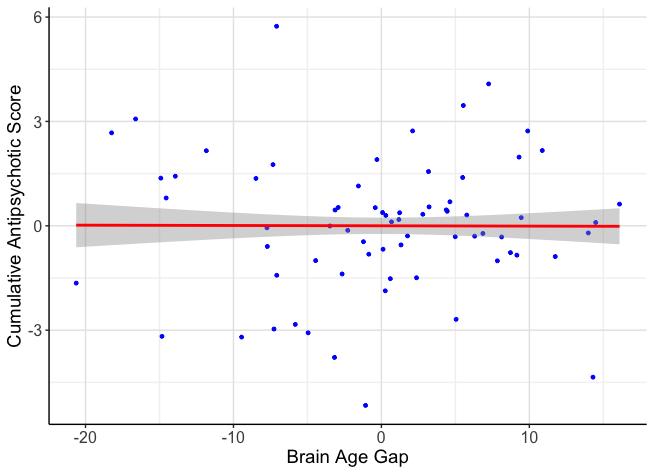

sFigure 5: Relationship Between Glutathione and PANSS Total Score

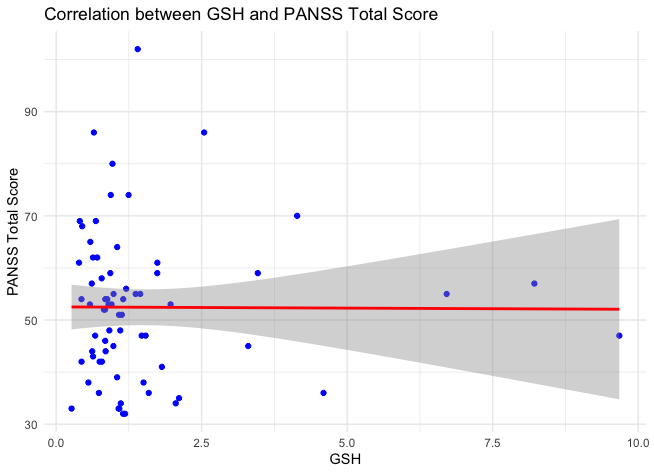

sFigure 6: Relationship Between Glutathione and Full Scale IQ

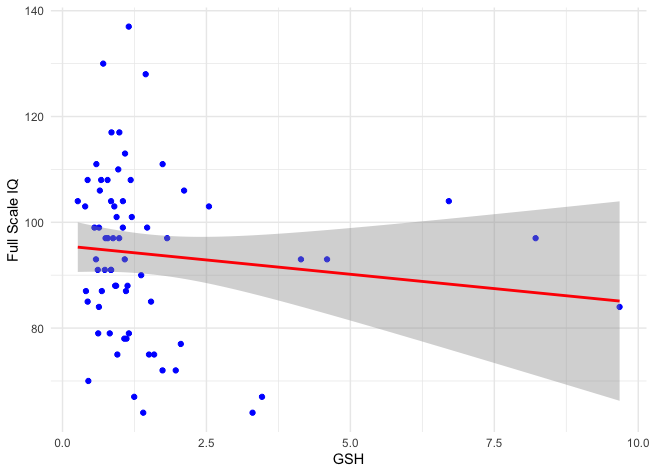

sFigure 7: Relationship Between Glutathione and Weschler Test of Adult Reading

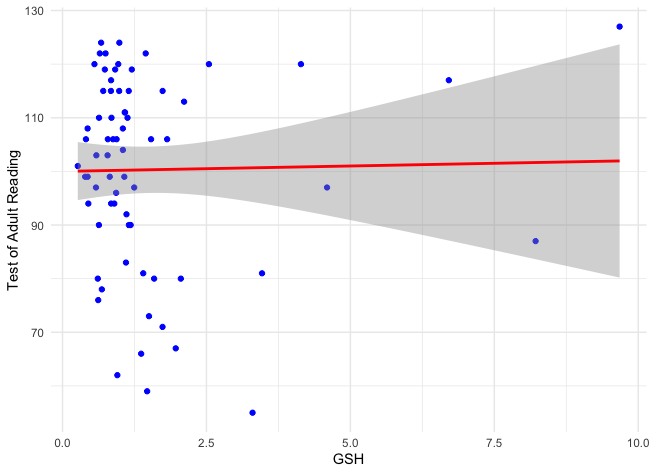

sFigure 3: Relationship Between BAG and Pro-inflammatory Factor 1 in total dataset

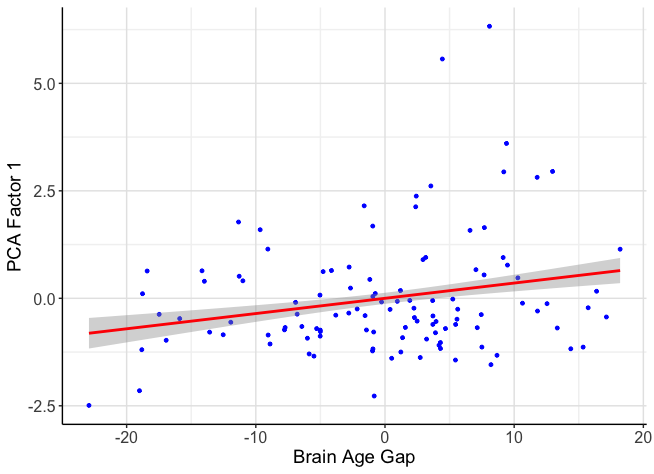

sFigure 9: Relationship Between Glutathione and Pro-inflammatory Factor 1

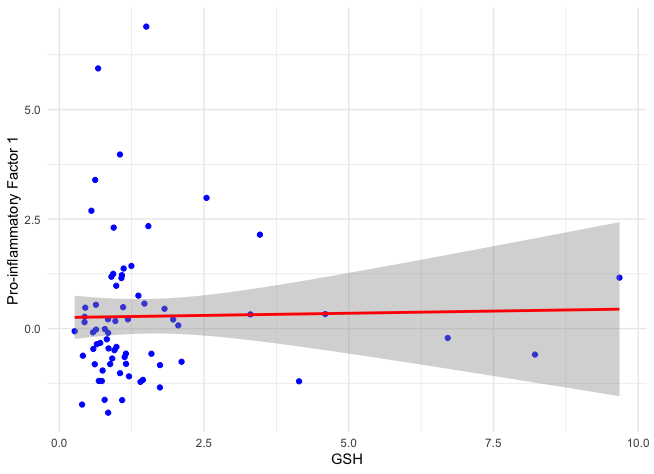

sFigure 10: Relationship Between Glutathione and Anti-inflammatory Factor 2

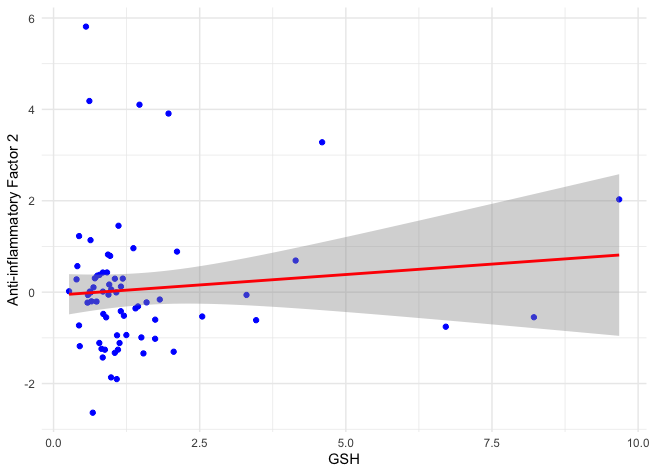

sFigure 11: Relationship Between Glutathione and Immuno-Modulation Factor 3

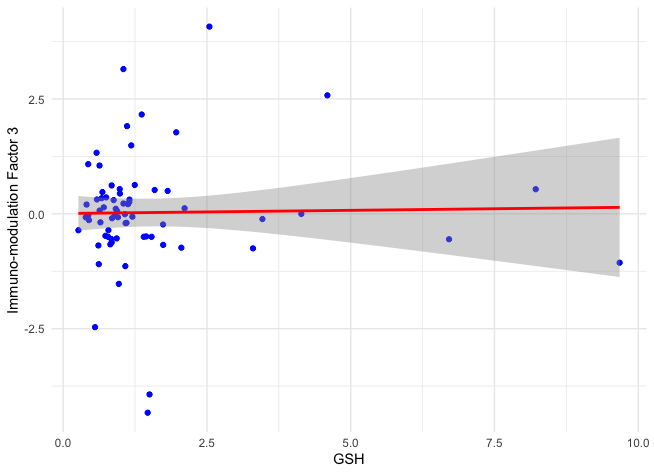

sFigure 12: Relationship Between Glutathione and T-Cell Modulation Factor 4

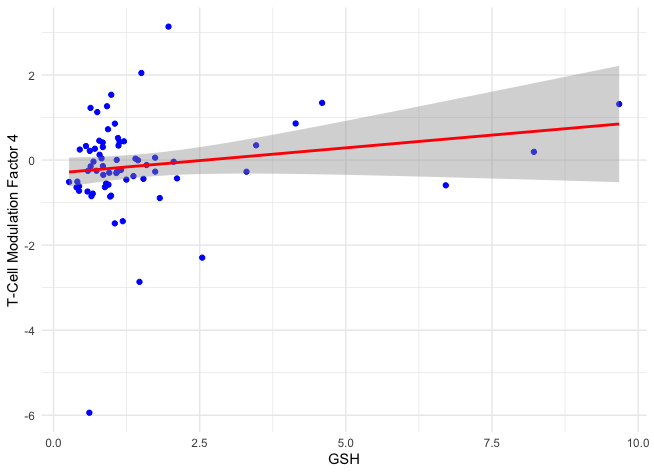

sFigure 13: Relationship Between Glutathione and Th1 Immune Response Factor 5

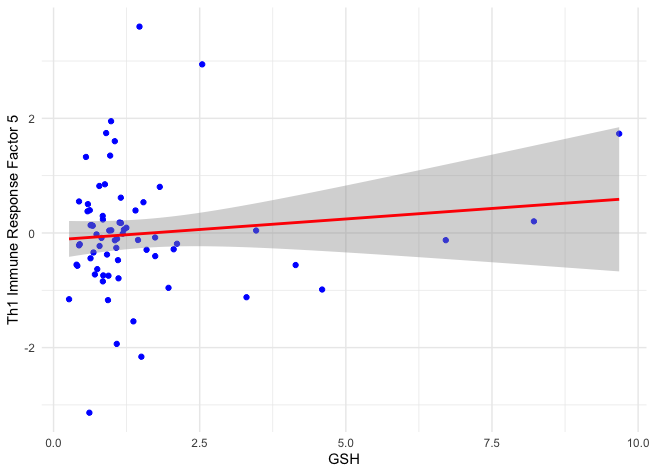
